# Racial and Sociodemographic Disparities in Blindness Associated with Primary Angle Closure Glaucoma in the United States: An IRIS^®^ Registry Analysis

**DOI:** 10.1101/2022.08.26.22279190

**Authors:** Sona N. Shah, Sarah Zhou, Carina Sanvicente, Bruce Burkemper, Galo Apolo, Charles Li, Siying Li, Lynn Liu, Flora Lum, Sasan Moghimi, Benjamin Xu

## Abstract

**Purpose:** To assess the prevalence and risk factors of blindness among patients newly diagnosed with primary angle closure glaucoma (PACG) in the United States (US).

**Design:** Retrospective cross-sectional study of patients from the American Academy of Ophthalmology IRIS^®^ (Intelligent Research in Sight) Registry.

**Participants:** Patients in the IRIS^®^ Registry between the years 2015 to 2019 with a new diagnosis of PACG and visual acuity (VA) data on or within 90 days prior to the date of diagnosis.

**Methods:** Eligible patients were aged 18 years and older and: (1) were observable in the database at least 24 months prior to the index date of PACG diagnosis; (2) had no history of intraocular pressure (IOP) lowering drops, laser peripheral iridotomy (LPI), cataract surgery, or a diagnosis of pseudophakia unless preceded by a diagnosis of anatomical narrow angle (ANA); and (3) had no history of glaucoma surgery. Multivariable logistic regression models were developed to assess risk factors of blindness.

**Main Outcome Measures:** Any (one or both eyes) or bilateral (both eyes) blindness (VA ≤ 20/200) at first diagnosis of PACG.

**Results:** 43,901 patients with PACG in the IRIS^®^ Registry met inclusion criteria. Overall prevalence of any and bilateral blindness were 11.5% and 1.8%, respectively. Black and Hispanic patients were at higher risk of any (OR=1.42 and 1.21, respectively; p<0.001) and bilateral (OR=2.04 and 1.53, respectively; p<0.001) blindness compared to non-Hispanic White patients adjusted for ocular comorbidities, including cataracts. Other factors associated with any blindness included age <50 or >80 years, male sex, Medicaid or Medicare insurance category, and Southern or Western practice region (ORs>1.28; p≤0.01). Diagnosis of ANA prior to diagnosis of PACG was protective against any (OR=0.56; p<0.001) and bilateral (OR=0.61; p<0.001) blindness.

**Conclusions:** Blindness affects 1 out of 9 patients with newly diagnosed PACG in the IRIS^®^ Registry; Black and Hispanic patients and Medicaid and Medicare recipients are significantly more vulnerable. These findings highlight the severe ocular morbidity associated with PACG and the need for increased disease awareness and improved detection methods.

## Introduction

Primary angle closure glaucoma (PACG) is a visually devastating disease and leading cause of irreversible blindness worldwide.^1-3^ The current global prevalence of PACG is approximately 23 million, although this number is projected to rise to approximately 32 million by 2040 due to the aging of the world’s population.^1^ Any rise in the prevalence of PACG is problematic due to high rates of blindness associated with the disease; it is estimated that blindness affects 27.0% of those with PACG worldwide.^3^ Quality of life (QoL), defined as an individual’s experience of health, comfort, happiness, and ability to enjoy day-to-day life activities, is significantly diminished by blindness, especially in bilateral cases.^4-6^ Blindness also has a profound impact on healthcare expenditures and the global economy; the economic burden of vision loss has been estimated to be $134.2 billion annually in the United States (US) alone.^7^ Therefore, there exists an urgent need to better understand the burden and risk factors of blindness secondary to treatable ocular diseases, such as PACG, on diverse patient populations.

While primary open angle closure glaucoma (POAG) is twice as common as PACG, PACG confers a two-fold higher risk of blindness.^3^ Therefore, POAG and PACG are associated with a similar number of cases of blindness worldwide.^8^ The visually devastating nature of PACG stems from anatomical mechanisms underlying the disease. Angle closure is characterized by contact between the peripheral iris and trabecular meshwork, which impedes outflow of aqueous humor from the eye.^9^ Extensive angle closure can lead to complete obstruction of outflow, elevated intraocular pressure (IOP) that is higher than typically observed in open angle eyes, and rapid glaucomatous damage to the optic nerve. Laser and surgical treatments help alleviate angle closure, which effectively delay or prevent the onset of elevated IOP and PACG.^10,11^ Therefore, many cases of irreversible blindness associated with PACG could be avoided if high-risk patients are identified and treated earlier in the disease course.

There are an estimated 700,000 people with PACG in the US.^1^ However, the visual impact of PACG in the US is poorly studied, due in part to the low perceived impact of PACG among non-Asian individuals and the racial and ethnic diversity of the US population. In this study, we use data from the IRIS^®^ Registry (Intelligent Research in Sight) to study the prevalence of blindness among patients newly diagnosed with PACG in the US. The US has the highest total health expenditure per capita of worldwide; therefore, it is reasonable to speculate that there should be lower ocular morbidity associated with PACG in the US compared to other regions of the world.^12-19^ We also assess sociodemographic factors to identify populations more vulnerable to PACG-associated blindness who could benefit from increased provider awareness and improved detection methods.

## Methods

### Data

The American Academy of Ophthalmology IRIS^®^ Registry is a comprehensive clinical registry that includes data on approximately 441 million patient visits for over 73 million unique patients as of April 1, 2022. Available eye-level clinical data (with specified laterality in the IRIS^®^ Registry) included dates of clinical diagnoses (including PACG), visual acuity (VA), intraocular pressure (IOP), and dates of procedures including laser peripheral iridotomy (LPI) and cataract and glaucoma surgeries. Available patient-level clinical data included history of IOP-lowering medications (laterality not specified in the IRIS^®^ Registry). Available patient-level sociodemographic data included age, race, sex, insurance category, and practice region. Race and ethnicity, which are cultural constructs with biological contribution through genetic heritage, are combined into a single variable in the IRIS^®^ Registry. This study was approved by The University of Southern California Institutional Review Board. The study adhered to the tenets of the Declaration of Helsinki and complied with the Health Insurance Portability and Accountability Act.

### Study Population Selection and Definitions

Inclusion in the study population required a new diagnosis of PACG. Eligible patients with newly diagnosed PACG were identified from the IRIS^®^ Registry between 2015 to 2019. Inclusion in the study population required an index diagnosis of PACG based on *International Classification of Diseases Ninth Revision* (ICD-9) or *Tenth Revision* (ICD-10) codes (Supplementary Table 1). PACG diagnoses were analyzed on the eye level. The index date of diagnosis was defined as the date of the first visit associated with a PACG diagnosis.

Eligible patients were aged 18 years and older with newly diagnosed PACG, defined as: (1) observable in the IRIS^®^ Registry for at least 24 months prior to the index date of PACG diagnosis; (2) no history of IOP lowering drops, LPI, cataract surgery, or a diagnosis of pseudophakia unless preceded by a diagnosis of ANA (also referred to as primary angle closure without glaucoma) based on ICD or Current Procedural Terminology (CPT) codes (Supplementary Table 1); (3) no history of glaucoma surgery (defined as trabeculectomy, glaucoma shunt, and cyclophotocoagulation) based on CPT codes (Supplementary Table 1). Criterion 1 was implemented to ensure that cases of PACG were newly diagnosed, based on the standard-of-care practice of monitoring PACG patients at least once per year. Criterion 2 was implemented to ensure that patients who had potentially received angle closure interventions for a diagnosis of ANA rather than PACG would not be excluded from the analyses. Criterion 3 was implemented to ensure that patients who had previously received glaucoma interventions would not be designated as newly diagnosed PACG.

Visual acuity data from the index date of diagnosis were analyzed on the eye level. Blindness was defined as VA ≤ 20/200 in at least one eye (any blindness) or in both eyes (bilateral blindness) on or within 90 days prior to the index date of diagnosis. If multiple VAs were documented within that time frame, the VA closest to the index date was utilized. Patients without VA data from at least one eye with PACG on or within 90 days prior to the index date of diagnosis were excluded from the analysis. Only patients with bilateral VA data were eligible for analyses on bilateral blindness.

### Statistical Analysis

Continuous data were expressed as means and standard deviations. Categorical data were expressed in proportions and percentages. Univariable and multivariable logistic regression analyses were performed to determine odds ratios (OR) for any and bilateral blindness adjusted for ocular comorbidities (cataracts, diabetic retinopathy, and macular degeneration diagnoses based on ICD codes). Variables significant at p < 0.15 in univariable analysis were included in multivariable analysis. Separate multivariable models were developed for any blindness, bilateral blindness, and any blindness additionally adjusted for IOP. The threshold for statistical and clinical significance was set at OR ≥ 1.20 and *p* ≤ 0.01 to avoid interpretation of weak effects due to large sample size. All statistical analyses were performed using R (The R Foundation for Statistical Computing, Vienna, Austria).

## Results

A total of 223,029 unique patients with an eye-level diagnosis of PACG from 2015 to 2019 were identified in the IRIS^®^ Registry (Figure 1). There were 43,901 unique patients who met criteria for newly diagnosed PACG after patients were excluded based on the lack of a minimum 24-month lookback period (116,535 patients excluded), presence of prior treatment and surgical history (48,326 patients excluded), or the absence of VA data from at least one eye with PACG on or within the 90 days prior to the index date of diagnosis (14,267 patients excluded).

**Figure 1.**
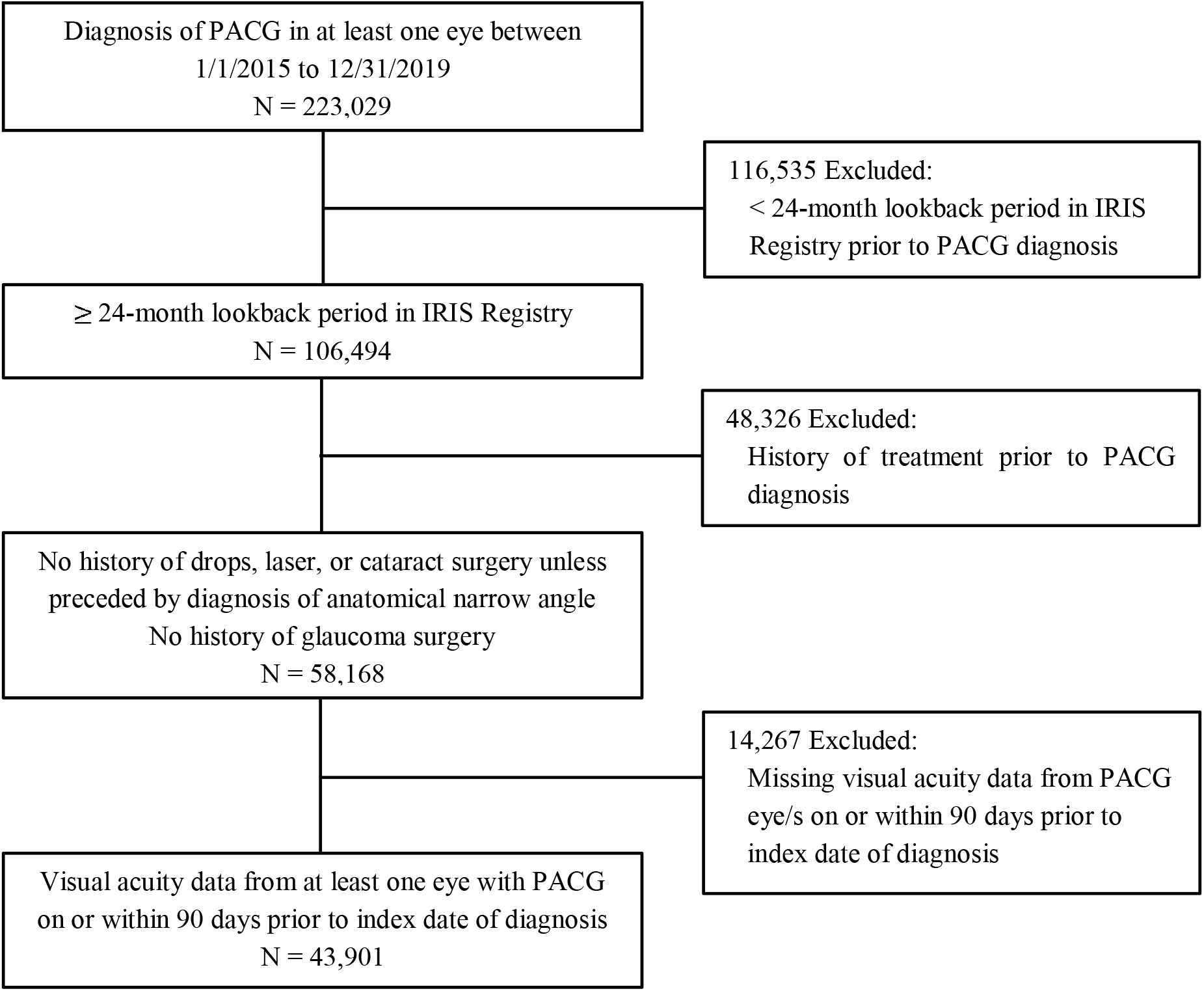
Attrition diagram of patients with newly diagnosed PACG in the IRIS® (Intelligent Research in Sight) Registry.

Out of 43,901 patients with VA data for at least one eye with newly diagnosed PACG, 5,064 (11.54%) were blind in at least one eye; out of 41,904 patients with bilateral VA data, 736 (1.76%) were blind in both eyes (Table 1). The effects of age and race on prevalence of any blindness and bilateral blindness were similar. The proportion of any and bilateral blindness were highest in patients < 40 (31.02% and 9.52%, respectively) and > 80 (18.60% and 3.60%, respectively) years of age; higher in males (13.71% and 1.95%, respectively) compared to females (10.41% and 1.66%, respectively); and higher in Black patients (14.2% and 2.93% respectively) and Hispanic patients (12.53% and 2.26%, respectively) compared to non-Hispanic White patients (Tables 1 and 2).

**Table 1.**
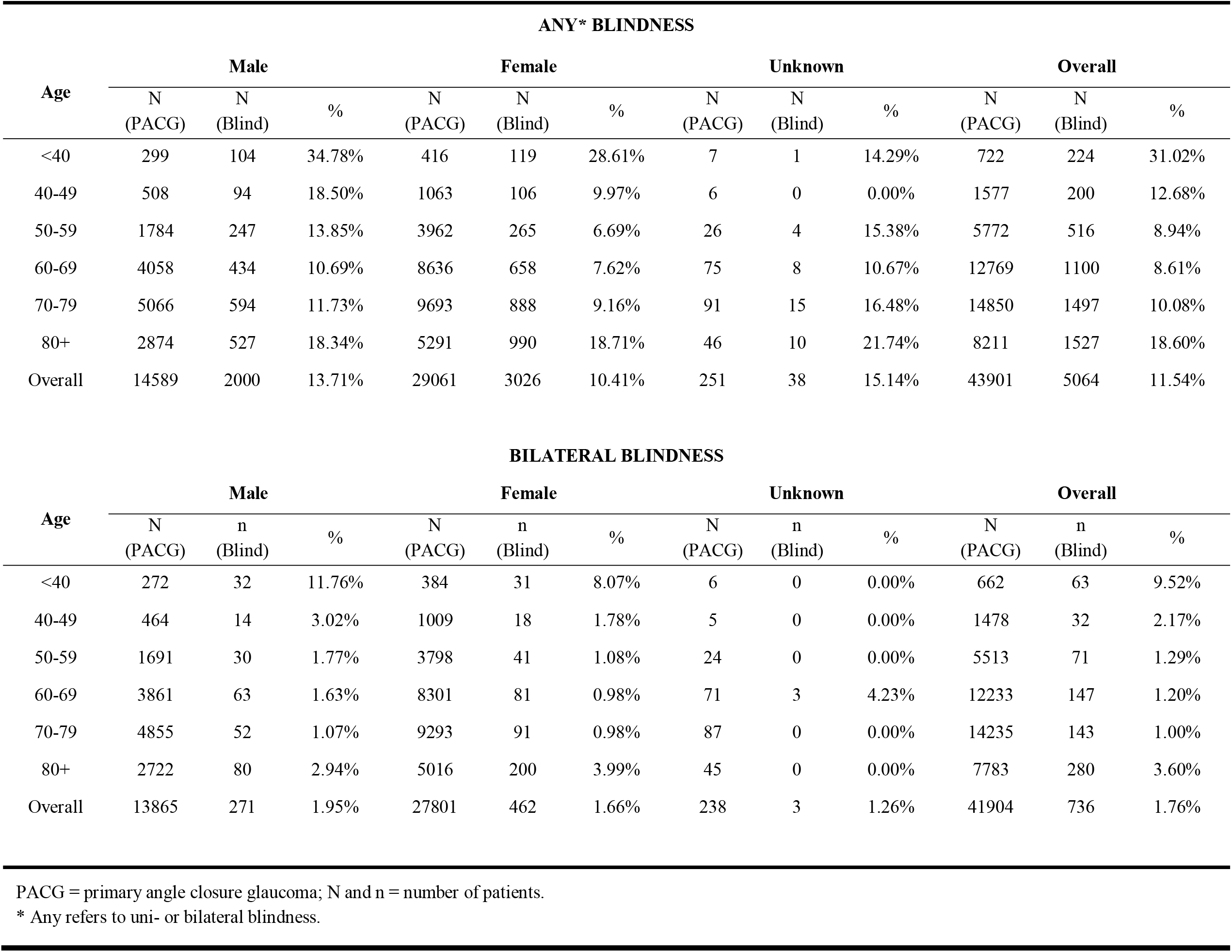
Proportion of blindness in newly diagnosed PACG cases stratified by sex and age.

**Table 2.** Proportion of blindness in newly diagnosed PACG cases stratified by race and age.

| ANY* BLINDNESS |  |  |  |  |  |  |  |  |  |  |  |  |  |  |  |  |  |  |
| --- | --- | --- | --- | --- | --- | --- | --- | --- | --- | --- | --- | --- | --- | --- | --- | --- | --- | --- |
| Age | Non-Hispanic White |  |  | Black |  |  | Hispanic |  |  | Asian |  |  | Unknown |  |  | Overall |  |  |
|  | N<br>(PACG) | n<br>(Blind) | % | N<br>(PACG) | n<br>(Blind) | % | N<br>(PACG) | n<br>(Blind) | % | N<br>(PACG) | n<br>(Blind) | % | N<br>(PACG) | n<br>(Blind) | % | N<br>(PACG) | n<br>(Blind) | % |
| <40 | 393 | 119 | 30.28% | 88 | 34 | 38.64% | 117 | 41 | 35.04% | 27 | 4 | 14.81% | 97 | 26 | 26.80% | 722 | 224 | 31.02% |
| 40-49 | 880 | 104 | 11.82% | 156 | 35 | 22.44% | 229 | 37 | 16.16% | 90 | 1 | 1.11% | 222 | 23 | 10.36% | 1577 | 200 | 12.68% |
| 50-59 | 3267 | 259 | 7.93% | 670 | 77 | 11.49% | 809 | 87 | 10.75% | 334 | 29 | 8.68% | 692 | 64 | 9.25% | 5772 | 516 | 8.94% |
| 60-69 | 7584 | 639 | 8.43% | 1471 | 155 | 10.54% | 1689 | 144 | 8.53% | 704 | 48 | 6.82% | 1321 | 114 | 8.63% | 12769 | 1100 | 8.61% |
| 70-79 | 9343 | 883 | 9.45% | 1422 | 193 | 13.57% | 1926 | 216 | 11.21% | 750 | 65 | 8.67% | 1409 | 140 | 9.94% | 14850 | 1497 | 10.08% |
| 80+ | 5610 | 1027 | 18.31% | 645 | 138 | 21.40% | 840 | 178 | 21.19% | 353 | 53 | 15.01% | 763 | 131 | 17.17% | 8211 | 1527 | 18.60% |
| Overall | 27077 | 3031 | 11.19% | 4452 | 632 | 14.20% | 5610 | 703 | 12.53% | 2258 | 200 | 8.86% | 4504 | 498 | 11.06% | 43901 | 5064 | 11.54% |

| BILATERAL BLINDNESS |  |  |  |  |  |  |  |  |  |  |  |  |  |  |  |  |  |  |
| --- | --- | --- | --- | --- | --- | --- | --- | --- | --- | --- | --- | --- | --- | --- | --- | --- | --- | --- |
| Age | Non-Hispanic White |  |  | Black |  |  | Hispanic |  |  | Asian |  |  | Unknown |  |  | Overall |  |  |
|  | N<br>(PACG) | n<br>(Blind) | % | N<br>(PACG) | n<br>(Blind) | % | N<br>(PACG) | n<br>(Blind) | % | N<br>(PACG) | n<br>(Blind) | % | N<br>(PACG) | n<br>(Blind) | % | N<br>(PACG) | n<br>(Blind) | % |
| <40 | 367 | 34 | 9.26% | 80 | 12 | 15.00% | 112 | 14 | 12.50% | 23 | 0 | 0.00% | 80 | 3 | 3.75% | 662 | 63 | 9.52% |
| 40-49 | 825 | 13 | 1.58% | 141 | 8 | 5.67% | 214 | 10 | 4.67% | 85 | 1 | 1.18% | 213 | 0 | 0.00% | 1478 | 32 | 2.17% |
| 50-59 | 3146 | 41 | 1.30% | 615 | 12 | 1.95% | 769 | 12 | 1.56% | 321 | 1 | 0.31% | 662 | 5 | 0.76% | 5513 | 71 | 1.29% |
| 60-69 | 7331 | 71 | 0.97% | 1376 | 26 | 1.89% | 1615 | 26 | 1.61% | 662 | 5 | 0.76% | 1249 | 19 | 1.52% | 12233 | 147 | 1.20% |
| 70-79 | 9001 | 64 | 0.71% | 1349 | 35 | 2.59% | 1844 | 25 | 1.36% | 693 | 7 | 1.01% | 1348 | 12 | 0.89% | 14235 | 143 | 1.00% |
| 80+ | 5333 | 180 | 3.38% | 606 | 29 | 4.79% | 795 | 34 | 4.28% | 331 | 12 | 3.63% | 718 | 25 | 3.48% | 7783 | 280 | 3.60% |
| Overall | 26003 | 403 | 1.55% | 4167 | 122 | 2.93% | 5349 | 121 | 2.26% | 2115 | 26 | 1.23% | 4270 | 64 | 0.10% | 41904 | 736 | 1.76% |

PACG = primary angle closure glaucoma; N and n = number of patients.
\* Any refers to uni- or bilateral blindness.

Multivariable logistic regression analysis showed significant associations between any blindness and the following parameters (Table 3): age groups < 40 (OR = 3.54, 95% CI 2.93-4.26), 40-49 (OR = 1.41, 95% CI 1.18-1.68), and ≥ 80 years (OR = 1.79, 95% CI 1.60-2.02) compared to 50-59 years; Black race (OR = 1.42, 95% CI 1.29-1.57) or Hispanic ethnicity (OR = 1.21, 95% CI 1.10-1.33) compared to non-Hispanic White race; Southern (OR = 1.34, 95% CI 1.23-1.46) or Western (OR = 1.31, 95% CI 1.19-1.44) practice regions compared to the Northeast practice region; Medicaid (OR = 2.06, 95% CI 1.75-2.42), Medicare fee-for-service (FFS) (OR = 1.51, 95% CI 1.37-1.65), or Medicare Managed (OR = 1.28, 95% CI 1.13-1.46) insurance category compared to private insurance; history of cataract (OR = 1.57, 95% CI 1.45-1.69) or macular degeneration (OR = 1.37, 95% CI 1.18-1.60); and PACG diagnosis without prior ANA diagnosis (OR =1.78, 95% CI 1.66-1.90). Female sex (OR = 0.75, 95% CI 0.71-0.80) was associated with significantly lower odds of any blindness compared to male sex.

**Table 3.**
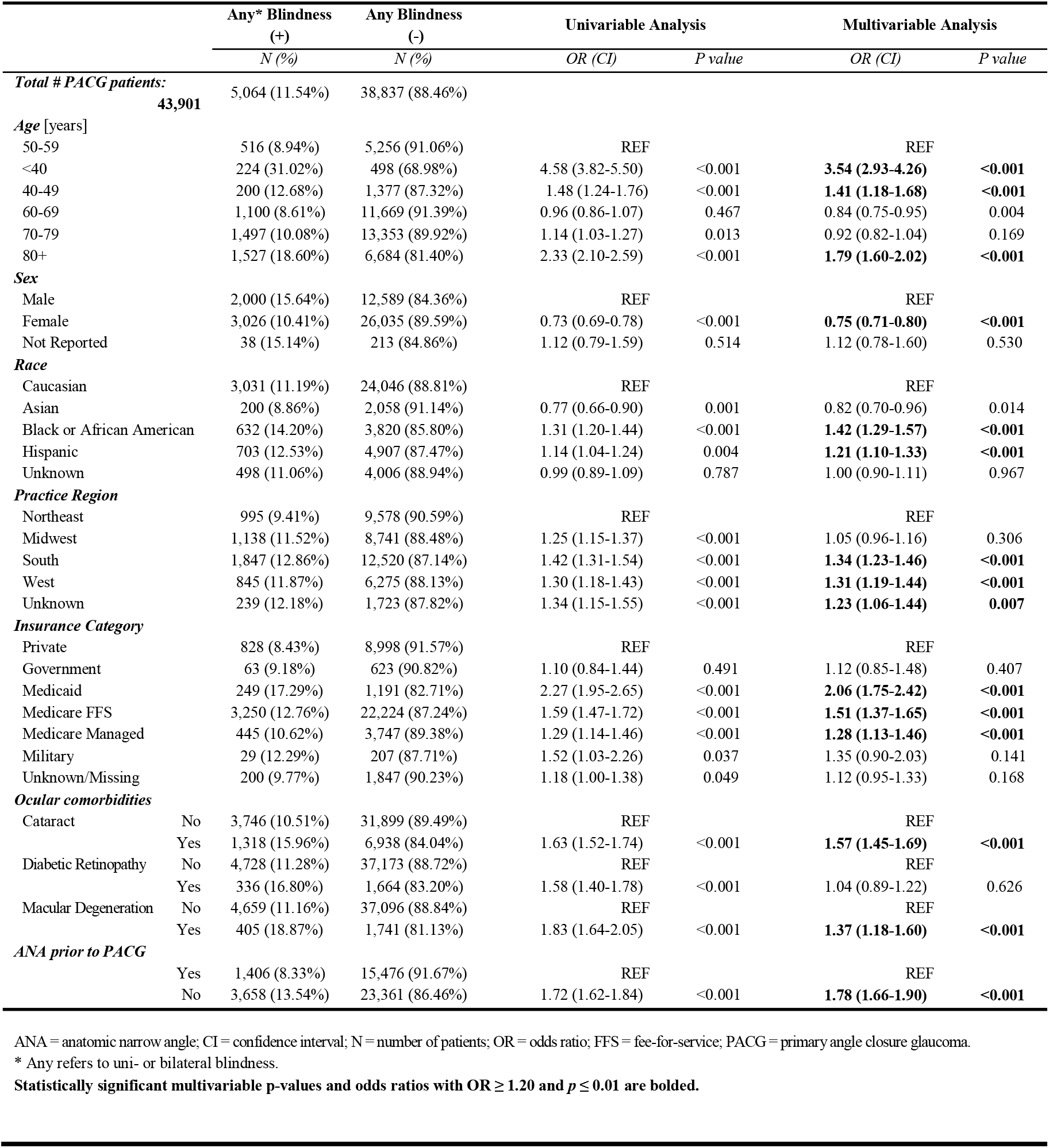
Univariable and multivariable analysis of risk factors for any* blindness in PACG.

Multivariable logistic regression analysis showed significant associations between bilateral blindness and the following parameters (Table 4): age groups < 40 (OR = 5.58, 95% CI 3.89-8.00) and > 80 (OR = 1.82, 95% CI 1.35-2.45) years of age compared to 50-59 years; Black race (OR = 2.04, 95% CI 1.64-2.53) or Hispanic ethnicity (OR = 1.53, 95% CI 1.23-1.90) compared to non-Hispanic White race; Southern (OR = 1.41, 95% CI 1.14-1.74) practice region compared to the Northeast practice region; Medicaid (OR = 3.85, 95% CI 2.71-5.48), Medicare fee-for-service (FFS) (OR = 2.70, 95% CI 2.07-3.51), or Medicare Managed (OR = 1.84, 95% CI 1.27-2.65) insurance categories compared to private insurance; history of cataract (OR = 1.48, 95% CI 1.23-1.78) or macular degeneration (OR = 1.74, 95% CI 1.25-2.40); and PACG diagnosis without prior ANA diagnosis (OR = 1.64, 95% CI 1.39-1.94) (Table 4). Age group 70-79 years (OR = 0.52, 95% CI 0.35-0.71) was significantly associated with lower odds of bilateral blindness compared to 50-59 years.

**Table 4.**
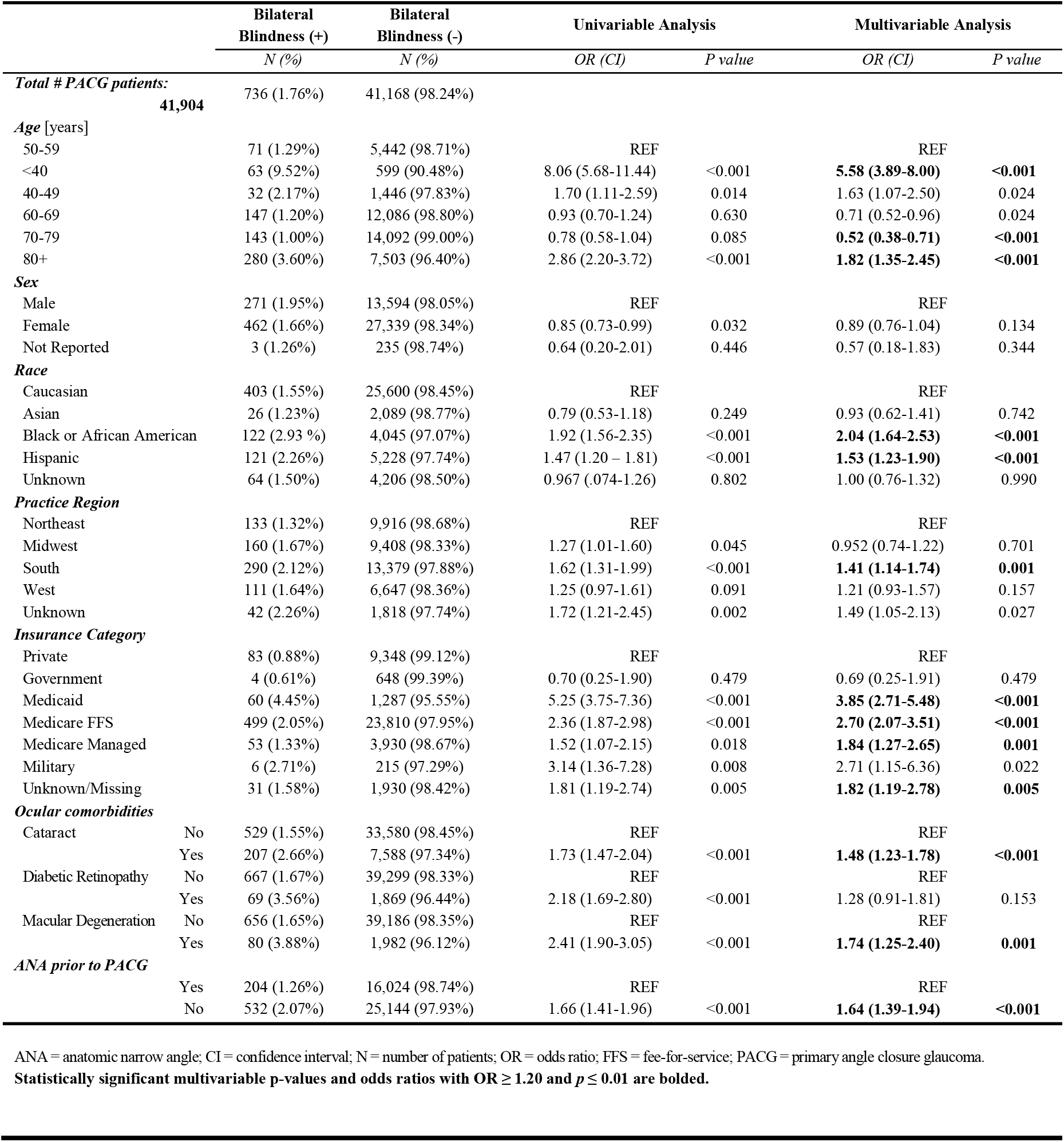
Univariable and multivariable analysis of risk factors for bilateral blindness in PACG.

The multivariable logistic regression model for any blindness was additionally adjusted for IOP (Supplementary Table 2). Results were similar to those from the multivariable model for any blindness without adjustment for IOP (Table 3); however, association with Western practice region was no longer significant, and diabetic retinopathy (OR = 2.00, 95% CI 1.65-2.42) was significantly associated with higher odds of any blindness (Supplementary Table 2).

## Discussion

In this study, we used data from the IRIS^®^ Registry to assess the prevalence and risk factors of blindness among patients with newly diagnosed PACG in the US. The overall prevalence of any and bilateral blindness among patients with newly diagnosed PACG were 11.5% and 1.8%, respectively. There were significant racial disparities in the prevalence of PACG-associated blindness; Black and Hispanic patients had 42% and 21% higher odds of blindness compared to non-Hispanic White patients. Other sociodemographic factors associated with blindness included age < 40 or > 80 years, male sex, Medicaid or Medicare insurance, and Southern or Western practice region. Diagnosis of ANA prior to diagnosis of PACG was protective against any and bilateral blindness. These findings highlight deficiencies in current practice patterns related to PACG; specifically, the need for increased disease awareness among eye care providers and development of more effective clinical methods to detect patients at high-risk of PACG.

The overall prevalence of any blindness among patients with newly diagnosed PACG in the US was relatively high regardless of age, sex, and race. It is well-established that PACG is a visually devastating disease: a meta-analysis of 23 population-based studies estimated the prevalence of PACG-associated blindness to be 27.0%.^3^ However, blindness rates vary widely by geographic region.^3,12,14-16,20,21^ For example, studies conducted in more developed countries and/or urban regions reported lower prevalence of blindness: 5.3% in the Tajimi Study, 6.1% in the Kumejima Study, and 10.2% in the Singapore Chinese Eye Study.^14,16,20^ Conversely, studies conducted in China consistently reported higher prevalence of blindness regardless of the degree of urbanization: 25.0% in the Beijing Eye Study, 25.5% in the Handan Eye Study, 42.9% in the Liwan Eye Study, and 71.7% in the Yunnan Minority Eye Study.^12,15,21,22^ However, prior to our study, there was sparse information on PACG-associated visual morbidity among racial and ethnic populations in the US. Our findings provide compelling evidence that the US does not have substantially lower rates of PACG-associated blindness compared to other developed countries despite having higher healthcare expenditure per capita.

There are significant racial and ethnic differences in the prevalence of PACG-associated blindness; in this study, Black (14.20%, 1 per 7.0 cases) and Hispanic patients (12.53%, 1 per 8.0 cases) were disproportionately affected compared to non-Hispanic White (11.19%, 1 per 8.9 cases) and Asian patients (8.86%, 1 per 11.2 cases). These racial disparities were magnified for bilateral blindness: 1 in 34.5 Black patients (2.93%) and 1 in 43.5 Hispanic patients (2.26%) were bilaterally blind compared to 1 in 64.5 non-Hispanic White patients (1.55%) and 1 in 81.3 Asian patients (1.23%). Furthermore, these disparities appear to be independent of IOP and ocular comorbidities, including cataracts. There are several likely explanations for these observations. First, it is widely recognized that PACG is most common among Asian individuals, with around half of global cases occurring in China.^8,16-18,23,24^ In contrast, there is sparse information about the prevalence of PACG among other races and ethnicities. The Beaver Dam Eye Study reported low prevalence of PACG among Black and non-Hispanic White participants, finding only two definite cases of narrow-angle glaucoma in 4,926 participants; however, it is unclear how often angle assessments were performed.^25^ Therefore, provider-level biases about racial or ethnic differences in PACG prevalence may contribute to greater vigilance about angle closure among Asian individuals. However, such biases should not lead to differences in angle closure detection between Black and non-Hispanic White patients, as both racial groups are thought to have low PACG prevalence. Second, there may be racial and ethnic differences in ocular biometry, such as anterior chamber depth, that could influence the likelihood of eye care providers performing gonioscopy to detect angle closure prior to development of glaucomatous damage.^26,27^ Finally, differences in utilization of and access to eye care services between racial and ethnic groups could influence visual outcomes.^28,29^ Overall, these findings support the need for additional research on racial and ethnic differences in anatomical mechanisms of angle closure and the development of more effective clinical methods to risk-stratify patients for PACG.

Older age over 80 years and younger age under 40 years were both risk factors for any and bilateral blindness compared to age 50-59 years. While age is a well-established risk factor for PACG, PACG tends to be rare in younger populations.^12,13,30,31^ We speculate that the anatomical mechanisms underlying PACG differ between younger and older patients affected by blindness.^31-33^ In older patients, cataract formation contributes to increased lens thickness and likely worsening of pupillary block and other lens-related mechanisms.^34^ In younger patients, PACG may be attributed to other mechanisms of angle closure, such as severe forms of plateau iris syndrome, nanopthalmos, or other developmental or congenital anomalies, that contribute to more advanced disease and profound vision loss at first diagnosis.^31,33^ Although PACG is rare in younger patients, more effective detection and treatment of PACG in this subpopulation would have a longer-lasting benefit for patients and healthcare systems.

Patient sex, practice region, and insurance category were identified as additional risk factors for blindness. Male sex was identified as a risk factor for any blindness, which could be attributed to lower utilization of medical care among men, leading to later disease detection.^35^ Patients seeking care in Western and Southern regions compared to the Northeast region were at higher risk for any and bilateral blindness, which could be attributed to differences in regional practices and screening methods. There may also be regional differences in access to care and density of eyecare providers that we could not account for due to limitations of IRIS^®^ Registry data.^36^ Finally, patients with Medicaid, Medicare FFS, or Medicare were at higher risk for any blindness compared to patients with private insurance. We speculate that insurance serves as a proxy for socioeconomic status in our analyses, as the IRIS^®^ Registry does not contain data on patient income, net worth, education level, or health literacy. Therefore, the increased risk of blindness associated with certain insurance products may reflect the effects of these confounding factors.

The diagnosis of ANA prior to the diagnosis of PACG was protective against any and bilateral blindness, suggesting that earlier detection of ANA is associated with more favorable clinical outcomes. Gonioscopy is the current clinical standard for detecting angle closure and distinguishing POAG from PACG.^2,37-39^ However, gonioscopy has a number of limitations, such as being expertise-dependent and potentially time-consuming or uncomfortable, that contribute to its underutilization.^38^ In addition, around three-quarters of patients with newly diagnosed PACG in the US do not have a prior diagnosis of ANA.^39^ While there are automated non-contact methods utilizing anterior segment OCT (AS-OCT) imaging and artificial intelligence (AI) to detect gonioscopic angle closure, these methods are not widely available for clinical use.^40-42^ Our findings on PACG-associated blindness highlight the need to further develop and implement more convenient clinical tools to detect patients at risk for PACG.

Our study has several limitations. First, our analyses relied on clinical diagnoses of PACG provided by a large number of practicing ophthalmologists, some of which may not comply with the formal definition of PACG proposed for epidemiological studies.^43^ It is feasible that some cases of PACG were misclassified cases of ANA (angle closure without glaucoma) or POAG; this would likely lower our estimates of PACG-associated blindness. However, we intentionally avoided applying additional criteria to narrow the definition of PACG, which could introduce systematic biases toward higher estimates of PACG-associated blindness. Second, we did not have access to visual field data. Therefore, we could not adopt a more comprehensive definition of blindness that includes <20 degrees of visual field; this would likely lead to further underestimation of the visual impact of PACG. Third, the duration of our lookback period to establish PACG cases as newly diagnosed was at minimum two years. As the IRIS^®^ Registry was recently launched in 2014, this substantially reduced our study sample size, which could limit the generalizability of our findings. Fourth, while we accounted for the three most common ocular co-morbidities (cataracts, age-related macular degeneration, and diabetic retinopathy) in our analyses, it is possible that other ocular diseases contributed to blindness. However, we believe that confounding is unlikely given that PACG has no known associations with systemic or other ocular diseases aside from cataracts. Finally, we cannot rule out that underlying racial and ethnic disparities in ocular health amplified racial and ethnic differences in the prevalence of blindness in our study.^44^

Our findings support that PACG is a visually devastating disease, even in the US. Historical epidemiological studies on PACG have shaped perception that the burden of PACG falls primarily on Asian patients, yet Black and Hispanic patients have significantly higher risk of PACG-associated blindness. These findings highlight the importance of renewed efforts to increase awareness about PACG and associated ocular morbidity. They also call into question the utilization of healthcare resources and the effectiveness of current practice patterns for detecting and managing patients at risk for PACG. We propose the development of more convenient and precise clinical tools for detecting and evaluating patients with angle closure, especially given the projected rise in PACG prevalence worldwide. Finally, this study demonstrates the importance and benefit of the IRIS^®^ Registry for studying the broader impact of ocular diseases within the US and mitigating their damaging effects by identifying and raising awareness about vulnerable patient populations.

## Data Availability

Data is not available

## Table and Figure Captions

Supplementary Table 1. Diagnosis, procedure, and treatment codes used in the study.

Supplementary Table 2. Univariable and multivariable analysis of risk factors for any blindness in PACG additionally adjusted for IOP.

## Acknowledgements

This work was supported by grants K23 EY029763 from the National Eye Institute, National Institute of Health, Bethesda, Maryland; an IRIS Registry Initiative Award from the American Glaucoma Society; and an unrestricted grant to the Department of Ophthalmology from Research to Prevent Blindness, New York, NY.

