## Supplementary Table 1 for "Racial and Sociodemographic Disparities in Blindness Associated with Primary Angle Closure Glaucoma in the United States: An IRIS^®^ Registry Analysis"

| **Supplementary Table 1.**  Diagnosis, procedure, and treatment codes used in the study. | | |
| --- | --- | --- |
| **Diagnosis Description** | ***ICD9*** | ***ICD10*** |
| Angle closure with glaucoma | 365.2* | H40.2* |
| Anatomical narrow angle, angle closure without glaucoma | 365.02, 365.06 | H40.03*, H40.069 |
| Age-related cataract | 366.* | H25.* |
| Pseudophakia | V43.1 | Z96.1 |
| **In-office Procedures** | ***CPT*** |  |
| Laser peripheral iridotomy | 66761 |  |
| **Glaucoma Surgeries** | ***CPT*** |  |
| Lens extraction / Cataract surgery | 66850, 66852, 66920, 66930, 66940, 66982, 66983, 66984 | |
| Trabeculectomy | 66170, 66172 |  |
| Aqueous shunt with or without graft | 66179, 66180 |  |
| Cyclophotocoagulation | 66710, 66711 |  |
| ICD9 = International Classification of Diseases, Ninth Revision; ICD10 = International Classification of Diseases, Tenth Revision; CPT = Current Procedural Terminology. | | |
