## Supplementary Table 2 for "Racial and Sociodemographic Disparities in Blindness Associated with Primary Angle Closure Glaucoma in the United States: An IRIS^®^ Registry Analysis"

| **Supplementary Table 2.** Univariable and multivariable analysis of risk factors for any* blindness in PACG additionally adjusted for IOP. | | | | | | | | | | | | |
| --- | --- | --- | --- | --- | --- | --- | --- | --- | --- | --- | --- | --- |
|  | | | **Any* Blindness**  **(+)** | **Any Blindness**  **(-)** |  | **Univariable Analysis** | | |  | **Multivariable Analysis** | | |
|  | | | *N (%)* | *N (%)* |  | *OR (CI)* | *P value* | |  | *OR (CI)* | *P value* | |
| ***Total # PACG patients:***  **43,901** | | | 5,064 (11.54%) | 38,837 (88.46%) |  |  |  | |  |  |  | |
| ***Age*** [years] | | |  |  |  |  |  | |  |  |  | |
| 50-59 | | | 516 (8.94%) | 5,256 (91.06%) |  | REF | | |  | REF | | |
| <40 | | | 224 (31.02%) | 498 (68.98%) |  | 4.58 (3.82-5.50) | <0.001 | |  | **3.66 (2.88-4.65)** | **<0.001** | |
| 40-49 | | | 200 (12.68%) | 1,377 (87.32%) |  | 1.48 (1.24-1.76) | | <0.001 |  | **1.49 (1.20-1.85)** | | **<0.001** |
| 60-69 | | | 1,100 (8.61%) | 11,669 (91.39%) |  | 0.96 (0.86-1.07) | 0.467 | |  | 0.99 (0.86-1.15) | 0.917 | |
| 70-79 | | | 1,497 (10.08%) | 13,353 (89.92%) |  | 1.14 (1.03-1.27) | 0.013 | |  | 1.12 (0.97-1.30) | 0.126 | |
| 80+ | | | 1,527 (18.60%) | 6,684 (81.40%) |  | 2.33 (2.09-2.59) | <0.001 | |  | 2.30 (1.98-2.67) | **<0.001** | |
| ***Sex*** | | |  |  |  |  |  | |  |  |  | |
| Male | | | 2,000 (13.71%) | 12,589 (86.29%) |  | REF | | |  | REF | | |
| Female | | | 3,026 (10.41%) | 26,035 (89.59%) |  | 0.73 (0.69-0.78) | <0.001 | |  | **0.80 (0.74-0.86)** | **<0.001** | |
| Not Reported | | | 38 (15.14%) | 213 (84.86%) |  | 1.12 (0.79-1.59) | 0.514 | |  | 0.86 (0.54-1.36) | 0.517 | |
| ***Race*** | | |  |  |  |  |  | |  |  |  | |
| Caucasian | | | 3,031 (11.19%) | 24,046 (88.81%) |  | REF | | |  | REF | | |
| Asian | | | 200 (8.86%) | 2,058 (91.14%) |  | 0.77 (0.66-0.90) | 0.001 | |  | 0.89 (0.74-1.07) | 0.215 | |
| Black or African American | | | 632 (14.20%) | 3,820 (85.80%) |  | 1.31 (1.20-1.44) | <0.001 | |  | **1.31 (1.1561.47)** | **<0.001** | |
| Hispanic | | | 703 (12.53%) | 4,907 (87.47%) |  | 1.14 (1.04-1.24) | 0.004 | |  | **1.39 (1.25-1.55)** | **<0.001** | |
| Unknown | | | 498 (11.06 %) | 4,006 (88.94%) |  | 0.99 (0.89-1.09) | 0.787 | |  | 1.03 (0.91-1.16) | 0.691 | |
| ***IOP*** | | | N/A | N/A |  | 1.07 (1.05-1.07) | <0.001 | |  | 1.06 (1.06-1.07) | <0.001 | |
| ***Practice Region*** | | |  |  |  |  |  | |  |  |  | |
| Northeast | | | 995 (9.41%) | 9,578 (90.59%) |  | REF | | |  | REF | | |
| Midwest | | | 1,138 (11.52%) | 8,741 (88.48%) |  | 1.25 (1.15-1.37) | <0.001 | |  | 1.01 (0.90-1.14) | 0.858 | |
| South | | | 1,847 (12.86%) | 12,520 (87.14%) |  | 1.42 (1.31-1.54) | <0.001 | |  | 1.14 (1.05-1.28) | **0.004** | |
| West | | | 845 (11.87%) | 6,275 (88.13%) |  | 1.30 (1.18-1.43) | <0.001 | |  | 1.14 (1.01-1.28) | 0.029 | |
| Unknown | | | 239 (12.18%) | 1,723 (87.82%) |  | 1.34 (1.15-1.55) | <0.001 | |  | 1.16 (0.95-1.41) | 0.131 | |
| ***Insurance Category*** | | |  |  |  |  |  | |  |  |  | |
| Private | | | 828 (8.43%) | 8,998 (91.57%) |  | REF | | |  | REF | | |
| Government | | | 63 (9.18%) | 623 (90.82%) |  | 1.10 (0.84-1.44) | 0.491 | |  | 1.10 (0.80-1.50) | 0.572 | |
| Medicaid | | | 249 (17.29%) | 1,191 (82.71%) |  | 2.27 (1.95-2.65) | <0.001 | |  | **1.61 (1.32-1.96)** | **<0.001** | |
| Medicare FFS | | | 3,250 (12.76%) | 22,224 (87.24%) |  | 1.59 (1.47-1.72) | <0.001 | |  | **1.42 (1.27-1.59)** | **<0.001** | |
| Medicare Managed | | | 445 (10.62%) | 3,747 (89.38%) |  | 1.29 (1.14-1.46) | <0.001 | |  | 1.15 (0.98-1.34) | 0.086 | |
| Military | | | 29 (12.29%) | 207 (87.71%) |  | 1.52 (1.03-2.26) | 0.037 | |  | 1.50 (0.94-2.40) | 0.090 | |
| Unknown/Missing | | | 200 (9.77%) | 1,847 (90.23%) |  | 1.18 (1.00-1.38) | 0.049 | |  | 0.99 (0.80-1.22) | 0.899 | |
| ***Ocular comorbidities*** | | |  |  |  |  |  | |  |  |  | |
| Cataract | | No | 3,746 (10.51%) | 31,899 (89.49%) |  | REF | | |  | REF | | |
|  | | Yes | 1,318 (15.96%) | 6,938 (84.04%) |  | 1.63 (1.52-1.74) | <0.001 | |  | **1.52 (1.38-1.66)** | **<0.001** | |
| Diabetic Retinopathy | | No | 4,728 (11.28%) | 37,173 (88.72%) |  | REF | | |  | REF | | |
|  | | Yes | 336 (16.80%) | 1,664 (83.20%) |  | 1.58 (1.40-1.78) | <0.001 | |  | **2.00 (1.65-2.42)** | **<0.001** | |
| Macular Degeneration | | No | 4,659 (11.16%) | 37,096 (88.84%) |  | REF | | |  | REF | | |
|  | | Yes | 405 (18.87%) | 1,741 (81.13%) |  | 1.83 (1.64-2.05) | <0.001 | |  | **2.50 (2.11-2.96)** | **<0.001** | |
| ***ANA prior to PACG*** | |  |  |  |  |  | | |  |  | | |
|  | Yes | | 1,406 (8.33%) | 15,476 (91.67%) |  | REF | | |  | REF | | |
|  | No | | 3,658 (13.54%) | 23,361 (86.46%) |  | 1.72 (1.62-1.84) | <0.001 | |  | **1.63 (1.50-1.76)** | **<0.001** | |
| ANA = anatomic narrow angle; CI = confidence interval; N = number of patients; OR = odds ratio; FFS = fee-for-service; IOP = intraocular pressure; PACG = primary angle closure glaucoma.  * Any refers to uni- or bilateral blindness.  **Statistically significant multivariable p-values and odds ratios with OR ≥ 1.20 and *p* ≤ 0.01 are bolded.** | | | | | | | | | | | | |
